# Bidirectional Associations between Short or Long Sleep Duration and Cognitive Function: the China Health and Retirement Longitudinal Study

**DOI:** 10.1101/2021.01.23.21250015

**Authors:** Jianian Hua, Hongpeng Sun, Qi Fang

## Abstract

**IMPORTANCE:** The bidirectional association between sleep duration and cognitive function has not been conclusively demonstrated.

**OBJECTIVE:** To investigate the longitudinal association between sleep duration and cognitive function among middle-aged and elderly Chinese participants.

**Design, SETTING, AND PARTICIPANTS:** A national representative and prospective longitudinal study in China. 7984 participants aged 45 years and above were assessed at baseline between June 2011 and March 2012 (wave 1) and 2013 (wave 2), 2015 (wave 3) and 2018 (wave4).

**MAIN OUCOMES AND MEASURES:** Self-reported nighttime sleep duration was evaluated by interview. Cognitive function was evaluated via assessments of global cognition, which reflected the ability of episodic memory, visuospatial construction, calculation, orientation and attention.

**Results:** Regarding the 7984 participants in wave 4, the mean (SD) age was 64.7 (8.4), 3862 (48.4) were male, and 6453 (80.7) lived in rural area. There were 14981, 11768 (78.6%), 10192 (68.0%), 7984 (53.3%) participants in the four waves of the study, respectively. Latent growth models showed both sleep duration and global cognition worsen over time. Cross-lagged models indicated that long or short sleep duration in the previous wave was associated lower global cognition in the next wave (standardized β=-0.066; 95%CI: −0.073, −0.059; P<0.001; Wave 1 to 2), and lower global cognition in the previous wave was associated with long or short sleep duration in the next wave (standardized β=-0.106; 95%CI: −0.116, −0.096; P<0.001; Wave 1 to 2). Global cognition was probably the major driver in this reciprocal associations.

**CONCLUSIONS AND REVELANCE:** There were bidirectional associations between long or short sleep duration and cognitive function. Lower cognitive function had a stronger association with worse cognitive function than the reverse. A moderate sleep duration is always recommended. Moreover, attention should be paid on the declined cognition and cognitive therapy among older adults with short or long sleep duration.

## 1 Introduction

Dementia is a common burden for global health. Approximately 50 million people had dementia in 2019 worldwide, and the number would triple by 2050^1^. Care and treatment for patients with dementia bring heavy costs for families, health systems, and society^2,3^. Considering there is no effective strategies, it is of great importance to learn modifiable risk factors for dementia.

Cognitive function test is a key way to detect dementia. The impact of sleep duration on cognitive function was widely reported, by cross-sectional studies^4^, longitudinal studies^5,6^, and the mendelian randomization study^7^. Solid epidemiological evidence supports an unidirectional association that both short and long sleep durations are associated with lower cognitive function^6^. Only a few studies learned the effects of cognitive function on sleep duration, reporting people with Alzheimer disease (AD) or mild cognitive impairment (MCI) had a higher prevalence of sleep disturbance^8-11^ and different sleep architecture^12-14^. However, these studies seldom investigated nocturnal sleep duration, and was not population-based. Most importantly, studies are still lacking to learn the bidirectional associations between sleep duration and cognitive function. We do not know whether sleep duration is a risk factor or predictor for lower cognition, or sleep duration is the result of cognitive decline^15^.

This study used the China Health and Retirement Longitudinal Study (CHARLS), a national representative cohort, to learn how the two factors were associated with each other among middle-aged and elderly Chinese participants. Using multiple waves of CHARLS, we aimed to assess the trends for sleep duration and cognitive function over time, the reciprocal relationship, and the primary driving factor.

## 2 Results

At baseline, the mean age was 59.4±9.7 years. 47.9% of the participants were male (Table 1). 76.3% of the participants living in rural area. The mean global cognition score of all participants at baseline was 10.4±4.4. The sleep duration of most participants was between 4 to 10 hours. 8.0% of the individuals slept less than 4 hours, while 0.7% of the participants slept more than 10 hours. Those who completed follow up tended to be male and live in rural areas. More people slept less than 4 hours or more than 10 hours. The mean global cognition score decreased as times went by.

**Table 1.** Characteristics of study population at each wave

|  | Wave 1 | Wave 2 | Wave 3 | Wave 4 |
| --- | --- | --- | --- | --- |
|  | 14981 | 11768 | 10192 | 7984 |
| Age, mean (SD)<sup>a</sup> | 59.4 (9.7) | 60.8 (9.1) | 62.4 (8.8) | 64.7 (8.4) |
| Male, No. (%) | 7169 (47.9) | 5662 (48.1) | 5512 (48.2) | 3862 (48.4) |
| Living area, No. (%) |  |  |  |  |
| Rural | 11591 (76.3) | 9276 (51.9) | 7925 (80.1) | 6453 (80.7) |
| Urban | 3550 (23.7) | 2492 (21.0) | 2267 (19.9) | 1531 (19.3) |
| Global cognition score, mean (SD) | 10.4 (4.4) | 10.6 (4.3) | 10.2 (4.3) | 9.7 (4.9) |
| Sleep duration, No. (%) |  |  |  |  |
| 4-10 h | 13779 (91.3) | 10658 (90.6) | 9276(91.0) | 7080 (88.6) |
| < 4 h | 1191 (8.0) | 1028 (8.7) | 856 (8.4) | 810 (10.2) |
| > 10 h | 11 (0.7) | 82 (0.7) | 60 (0.6) | 94 (1.2) |

a: Age at the specific wave.

### 2.1 Latent Growth Models

First, we calculated the mean baseline value (intercept) and the rate of change (slope), for sleep duration and global cognition separately, using latent growth models (Table 2). The intercept of sleep duration was 0.071 (95%CI: 0.048, 0.094; P<0.001). The average worsening per year (slope) was 0.006 (95%CI: 0.006, 0.006; P<0.001). This model fitted excellent (CFI=0.989, TLI=0.987, RMSEA=0.018). The intercept of global cognition was 11.362 (95%CI: 11.321, 11.403; P<0.001). The decline rate per year (slope) was −0.201 (95%CI: −0.207, −0.195; P<0.001). This model fixed excellent (CFI=0.982, TLI=0.978, RMSEA=0.084). The variances of all variables were statistically significant, which indicated that there were individual differences in intercept and slope for sleep duration and global cognition.

**Table 2.** Latent growth models for sleep duration and global cognition separately^a^

| Trajectory | Intercept (SE) | Intercept Variance (SE) | Slope (SE) | Slope Variance (SE) |
| --- | --- | --- | --- | --- |
| Sleep duration | 0.071 (0.003)* | 0.023 (0.002)* | 0.006 (0.001)* | <0.001 (<0.001)* |
| Global cognition | 11.362 (0.041)* | 8.879 (0.224)* | -0.201(0.006)* | 0.028 (0.007)* |

a: 1 for sleep duration>10 or sleep duration<6; 0 for $6 \leq \text{sleep duration} \leq 8$ ;
\*: $P < 0.001$

**Table 3.** Cross-lagged models for all four waves*

|  | Wav1 1 to 2 | Wave 2 to 3 | Wave 3 to 4 |
| --- | --- | --- | --- |
| Standardized $\beta$ (SE) | | | |
| Sleep duration to global cognition | -0.066 (0.007) | -0.081(0.008) | -0.073(0.007) |
| Global cognition to sleep duration | -0.106(0.010) | -0.092(0.000) | -0.091(0.009) |
| Sleep duration to sleep duration | 0.580(0.011) | 0.006(0.014) | 0.708(0.016) |
| Global cognition to sleep duration | 0.624(0.008) | 0.807(0.008) | 0.675(0.007) |
| Unstandardized $\beta$ (SE) | | | |
| Sleep duration to global cognition | -0.249(0.026) | -0.249(0.026) | -0.249(0.026) |
| Global cognition to sleep duration | -0.034(0.003) | -0.034(0.003) | -0.034(0.003) |
| Sleep duration to sleep duration | 0.737(0.019) | 0.737(0.019) | 0.737(0.019) |
| Global cognition to sleep duration | 0.602(0.009) | 0.602(0.009) | 0.602(0.009) |

\*: $P < 0.001$ for all values.

Second, the intercept and slope of sleep duration and global cognition were considered together, using simultaneous equations (Figure S2). The overall model fit was excellent (CFI=0.985, TLI=0.974, RMSEA=0.034). The significant paths were drawn in solid lines, while the nonsignificant paths were drawn in dotted lines. The specific results were shown in Table S1. Global cognition at baseline correlated with the rate of change in global cognition (global cognition intercept-global cognition slope, r=0.267; 95%CI:0.239, 0.295; P<0.001). People with lower intercept tended to have a higher decline rate, and vice versa. Baseline sleep duration was associated with baseline global cognition (sleep duration intercept-global cognition intercept, r=-0.120, 95%CI: −0.129, −0.111; P<0.001). Which means too long or too short sleep duration was associated with lower global cognition cross-sectionally. Other four paths had relatively small P values (P<0.1). However, their clinical effects, reflected by r value, were limited. All covariates, except that sex was not associated with worsening rate of sleep duration (P=0.948), were statistically significantly associated with all four estimated parameters, including sleep duration intercept, sleep duration slope, global cognition intercept and global cognition slope (Table S2).

### 2.2 Cross-lagged models

The combined latent growth models could only reflect the existence and direction of the association. However, it could not determine whether the changes in global cognition were due to sleep duration, or the reverse. Therefore, we conducted cross-lagged models to explore the extent to which sleep duration and global cognition predicted one another over time (Figure 1). Paths constrained to be equal were marked with the same letter. The autoregressive paths (c and d, Figure 1) accounted for the stability of the measures. The cross-lagged paths (a and b) indicated the extent to which sleep duration and global cognition predicted one another over time.

**Figure 1.**
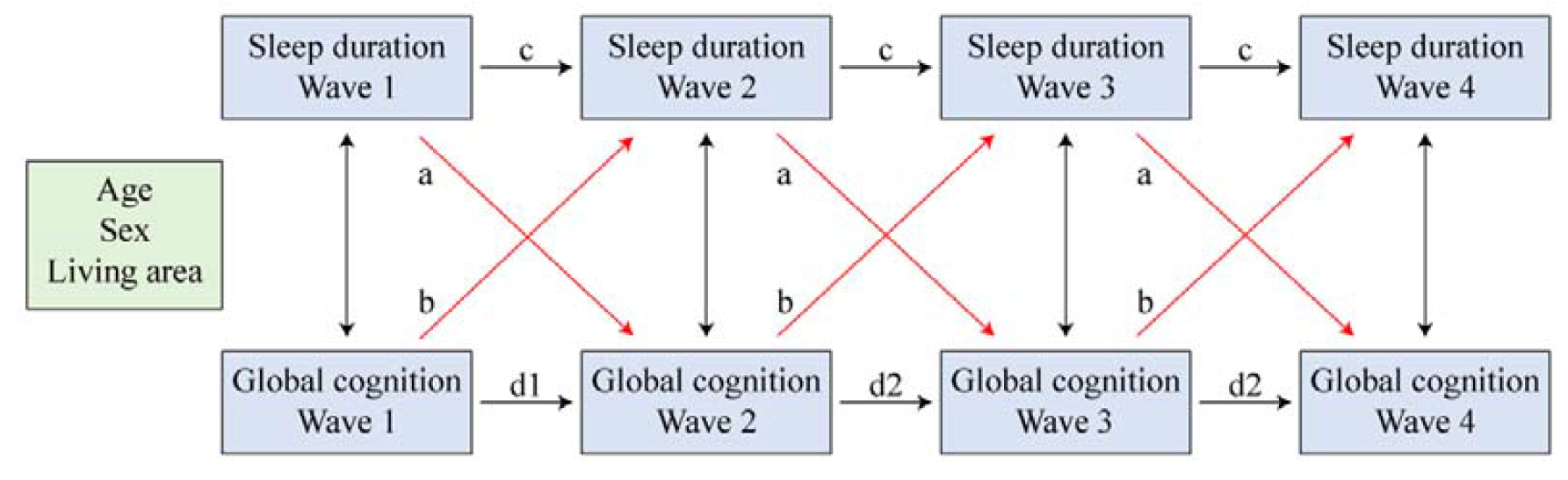
Cross-lagged models of sleep duration and global cognition.

The cross-lagged models including all four waves fitted good (CFI=0.962, AFI=0.906, RMSEA=0.059). The sleep duration in one wave was associated with sleep duration in the subsequent wave (unstandardized β=0.737; 95%CI: 0.718, 0.756; P<0.001); the global cognition in one wave was also associated with global cognition in the next wave (unstandardized β=0.602; 95%CI: 0.593, 0.611; P<0.001).

Sleep duration in the previous wave was associated with global cognition in the following wave (unstandardized β=-0.249; 95%CI: −0.275, −0.223; P<0.001); while global cognition in the previous wave was also associated with sleep duration in the following wave (unstandardized β=-0.034; 95%CI: −0.037, −0.031; P<0.001). Thus, there was a reciprocal association between sleep duration and global cognition during four waves. Results of the confounders was shown in Table S3.

While comparing the standardized beta coefficients, which is more comparable, the effect of global cognition on sleep duration (standardized β=-0.106; 95%CI: −0.116, −0.096; P<0.001; Wave 1 to 2) was larger than the reverse (standardized β=-0.066; 95%CI: −0.073, −0.059; P<0.001; Wave 1 to 2). This suggested that global cognition was the main driver in this reciprocal association.

### 2.3 Sensitivity analysis

The forest plot (Figure 2) compared the standardized beta coefficients of the two effects (sleep duration on global cognition and the reverse). The subgroups were based on different cut-points for optimal sleep duration. The pooled effect of global cognition on sleep duration was stronger.

**Figure 2.**
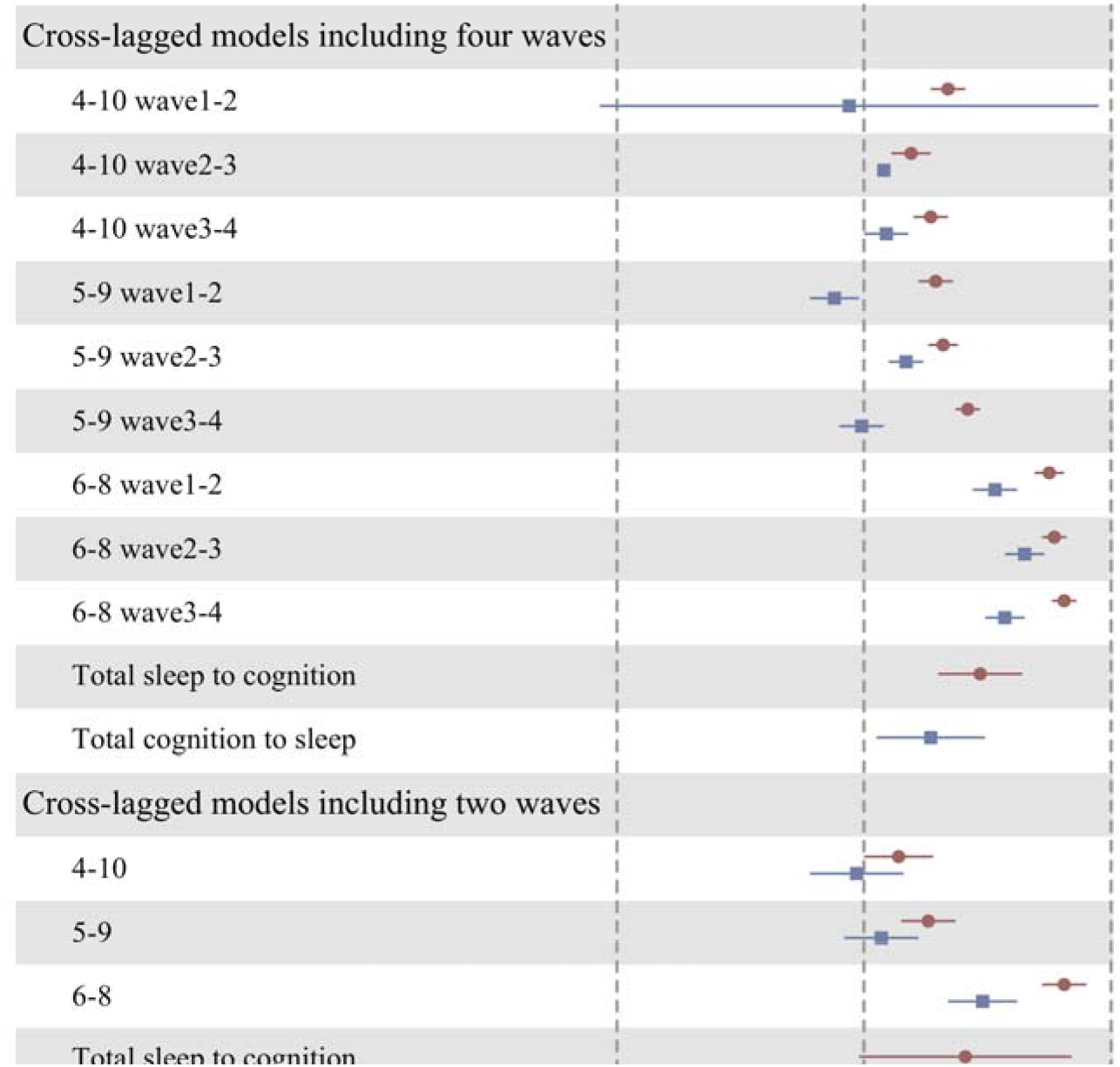
Forest plot based on different cu-off points and models. The red bars showed the effect of sleep duration on the previous wave to the global cognition on the next wave. The blue bars showed the effect of global cognition on the previous wave to the sleep duration on the next wave. “4-10”, “5-9”, and “6-8” meant different cut-off points.

The results were materially unchanged in the cross-lagged models including only Wave 1 and Wave 3. The sleep duration and global cognition in two waves were reciprocal associated with each other. The effect of global cognition on sleep duration was also stronger (Table S4, Figure 2).

## 3 Methods

### 3.1 Study population

The design of the CHARLS cohort has been described in detail elsewhere^16^. It was a community-based nationally representative longitudinal cohort study conducted in China, and a sister study of the Health and Retirement Study (HRS) in America. The Wave 1 (baseline) of the CHARLS took place in 2011, and included approximately 18000 randomly selected Chinese residents who were enrolled via multistage probability sampling. Follow-up surveys were conducted every two years. In the present study, we used data from Wave 1 (2011), Wave 2 (2013), Wave 3 (2015) and Wave 4 (2018) of the CHARLS.

15700 individuals had data on complete cognitive function tests and sleep duration in Wave 1. Because the subjects of the CHARLS were individuals aged ≥ 45 years, 344 individuals under 45 years old at baseline were excluded. 375 individuals with mental retardation at baseline were excluded. In the follow-up waves, people missing data on sleep duration or cognitive function were excluded^17^.Among the remaining 14981 participants, there were 11768 (78.6%) participants in Wave 2, 10192 (68.1%) participants in Wave 3, and 7984 (50.1%) participants in Wave 4 (Table 1).

### 3.2 Sleep duration

The sleep duration in this article was the nocturnal sleep duration, and in all four waves, it was assessed by asking participants the following question: “During the last month, how many hours of actual sleep did you get at night averagely”? The duration was accurate to 0.5 h.

In the main analysis, the sleep duration value of those who sleep less than 4 h or long than 10 h was defined as “1”, and the sleep duration value of those who slept between 4 h and 10 h was defined as “0”. The selection of the cut-off point was based on a previous study, reporting at most 4 or at least 10 hours of sleep duration could accelerate the cognitive decline duration a follow-up of 4 years, among Chinese^6^. The optimal sleep duration for cognitive function of our study population was approximately 7 hours. In the sensitivity analysis, we examined other two cut-off points: 5-9 h, and 6-8 h.

### 3.3 Cognitive Assessment

The cognitive assessment was conducted in all four waves and included three domains: episodic memory; visuospatial construction, as measured by figure drawing test; orientation, attention and calculation, as measured by TICS. The global cognition score, resulted from the sum of these three scores, represented participants’ cognitive function, and it was considered to be the primary outcome of interest. The global cognition score ranged from 0 to 21^18,19^.

The episodic memory test comprised immediate and delayed (five minutes later) recall for 10 unrelated words. The test score was the average number of immediate and delayed recall words and ranged from 0 to 10^20^. In the figure drawing test, which was used to assess visuospatial construction, the participants were shown a picture and asked to redraw it. One point was given for succeeding. The Telephone Interview of Cognitive Status (TICS) in CHARLS was based on selected questions in mini-mental status examination (MMSE)^21^, and comprised ten mental questions: a serial subtraction of 7 from 100 for five times, the date (day, month, year), the day of the week, and the season. It reflected participants’ abilities of attention, orientation and calculation. This score ranged from 0 to 10.

### 3.4 Covariates

The following covariates were included: baseline age (in years), sex, and baseline living area (urban and rural).

### 3.5 Sensitivity Analysis

While only including Wave 1 and Wave 3 in the cross-lagged models, we could reach a better model fitness (Supplementary Table 4). We concluded it to the following reasons. First, the global cognition in Wave 1 and Wave 3 had similar standard deviation (Table 1), which was more homogeneous. Second, the time interval between the two waves is longer, during which time more cognitive decline could be observed. Thus, we conducted cross-lagged model for the two waves, as a sensitivity analysis.

We further chose “5h-9h” and “6h-8h” as the cut-off point for moderate sleep duration, and then conducted cross-lagged models. We estimated the pooled beta coefficient and 95% CI, using random-effect models. Heterogeneity between the studies was assessed using Cochran Q statistics (P<0.1 indicates statistically significant heterogeneity) and I^2^ statistic (I^2^>50% indicates statistically significant heterogeneity)^22,23^.

We defined people with “excessive change” as those who slept too short at the previous wave and too long at the next wave, or the reverse. Our previous study found “excessive change” was strongly associated with lower cognitive function^4,24^ (Figure S1). Since we defined both short and long sleep duration as “1” in this article, “excessive change” could not be observed and cause bias. Fortunately, in the final database which included 7984 participants, none of the participants exhibited “reverse change”.

### 3.6 Statistical analysis

Both global cognition score and sleep duration would decline because of aging, which was reported by our previous study learning the CHARLS and articles learning other cohorts^4^. The kind of latent growth model we used was the latent linear growth model, in which the repeatedly measured scores for an individual were hypothesized to follow a line that can be described by an intercept (baseline) for each individual and a slope (rate of change) for each individual. As shown in Table 2, it provides a description of intraindividual/within-person mean for the repeated measure (intercept/baseline), interindividual/between-person variance in the intercept (intercept variance), intraindividual change (slope) and interindividual difference in intraindividual change (slope variance)^25^.

First, the latent growth models were used to examine the trends for sleep duration value and global cognition score, separately (Table 2)^26^. Second, the sleep duration model and the global cognition model were combined into one model (Figure S1). This combined model further examined the associations between sleep duration intercept, sleep duration slope, global cognition intercept, and global cognition slope, and covariates. In the combined model, the equations were estimated simultaneously while controlling for covariates.

In cross-lagged models, sleep duration at each time was regressed on sleep duration at the prior time and global cognition at the prior time. Similarly, global cognition at each time was regressed on sleep duration and global cognition of the prior time. Sleep duration and global cognition at the same time were correlated. All equations were estimated simultaneously while controlling for covariates. The standardized coefficients were in a common metric of standard error (SD). To achieve a more parsimonious model, we constrained the cross paths to be equal across time. We used weighted least squares estimator (WLSMV) that employs regression for categorical outcomes^27^.

Both latent growth models and cross-lagged models were examined using model fit statistics including the Comparative Fit Index (CFI), Tucker-Lewis Index (TLI), and root mean square error of approximation (RMSEA). CFI/TLI over 0.85 indicates good fit and values above 0.95 indicates excellent fit. We used a sandwich estimator for the standard errors that is robust to nonnormality^28^.

Statistical analyses were 2-sided, with α=.05 as the threshold for statistical significance. The pooled effects in the sensitivity analysis were calculated by Stata 15.1 (Stata Corp, College Station, TX). The forest plot was drawn by R-3.4.3. Other analyses were performed by SAS version 9.4 (SAS Institute Inc., Cary, NC, USA) and Mplus 8.3 (Muthén & Muthén) ^27,29^.

## 4 Discussion

Our study found not only short or long sleep duration was associated with lower cognition or cognitive decline, but also, lower cognitive function would bring with short or long sleep duration. More importantly, global cognition had a stronger influence on subsequent change in sleep duration than the reverse.

Based on the characteristics of study population at each wave and the latent growth models, the proportion of people with “too long” and “too short” sleep duration, over 90% of which were short sleepers, tended to be larger over time. This was due to the average sleep duration of population would decline during aging^4,30^. The cognitive function of our participants also declined over time. Meanwhile, people with lower baseline cognitive function tended to suffer from a faster decline. Since the sleep duration value was dichotomous, it was easy to understand there was no association between its intercept and slope. The sleep duration and cognitive function was associated with each other by “intercept-intercept”. This did not mean they were associated other only at baseline. They also correlated with each other cross-sectionally in any other waves, which has been widely recognized and reported by our previous study using CHARLS cohort^4^. The mean baseline of sleep duration was associated with the mean baseline of cognitive function. Changes in sleep duration was not affected by other variables. Comparing the effect of sleep duration at one wave on cognitive function at next wave vs the reverse, the standardized regression coefficient of global cognition to sleep duration value was larger.

Our findings from the latent growth models and cross-lagged models was partly in agreement with other studies. *Zheng 2018* reported intercept of cognitive function was correlated with intercept of visual impairment (r=-0.226), and the effect of visual impairment on cognitive function was twice as stronger as the reverse. However, they found a association between slope of cognitive function and slope of visual impairment (r=-0.139)^31^. *Stefan 2018* reported the effect of internet use on cognitive function was about 10 times as stronger as the reverse^27^. While *Lu 2020* found the effect of cognitive function was in dominance^32^, by examining the bidirectional association between edentulism and cognitive function, using two waves of CHARLS. Whether sleep was associated with cognitive decline was still controversial^6,15^. We did not find a significant association between decline rate of global cognition and sleep duration, in the combined latent growth models.

Several mechanisms could explain the impact of sleep duration on cognition. Short sleep duration correlated with many pathologies which would lead to lower cognition, such as impaired β-amyloid (Aβ) clearance, pathologic tau, impaired synaptic plasticity, and circadian rhythm disturbances^33-35^. Long sleep duration was associated with sleep fragmentation and chronic inflammatory, which were associated with lower cognition^36,37^.

Mechanisms explaining the effect of low cognitive function on sleep duration remained to be elucidated. An Italian study found people with AD or MCI had a higher risk of sleep disordered breathing, insomnia, REM behavior disorder, among 431 patients^9^. The sleep disturbance and sleep architecture mentioned above might decrease or increase sleep duration. Changes in sleep architecture might be contributed to AD pathology-induced neuronal and synaptic damage in sleep areas in the brain^10,38^. Decreasing cholinergic neurons in the basal forebrain, one of the sleep regulating centers, would change both cognition and sleep-wake cycle^39,40^. Lower cognition was associated with brain volume loss, which could decrease REM sleep duration^41,42^. AD patients could have dysfunctional circadian rhythms, which would change sleep duration^42,43^.

The foremost strength of our study was that we extended the previous research, which mostly focusing on the effect of sleep duration on cognitive function, to the bidirectional influence of one on the other. By using cross-lagged models, we firstly investigate the associations between two factors simultaneously and found cognitive function was the dominating factor. Other strength of our study included large number of participants, nationally representative cohort, and up-to-date data.

Our study also had several limitations. Firstly, the harmonized cognitive test in CHARLS was not a test used worldwide, such as MMSE. Some cognitive domains in MMSE were not measured, including naming, and complex commands^44^. Our cognitive tests might not reflect the real capacity of cognitive performance. However, our test is widely used and has a high correlation with MMSE^45^. Plenty of articles had proved its reliability and validity^46,47^. Declined memory is a risk factor for the development of dementia^48^. Our test had a higher share for episodic memory score, which is more suitable for research learning both sleep and cognition^48,49^.

Secondly, CHARLS recorded sleep duration in a subjective measurement, instead of objective measurement (polysomnography), and did not include other dimensions of sleep. Insomnia, daytime sleepiness, sleep quality, and sleep-disordered breathing were correlated with cognitive function. However, the difference between self-reported sleep duration and the objective was smaller among Chinese individual (49 min, 95% CI: 37-61), compared with that among other ethnics^50^. The subjective sleep duration was more utilized and applicable in epidemiologic studies. The cognitive function should also be considered by objective markers in the future, such as brain MR^51^.

Thirdly, both people with too short and too long sleep duration was defined as one category. It prevented us from distinguishing between short sleep duration and long sleep duration. About 90% of them were short sleepers. The effect of long sleep duration might be covered, which might lead to bias.

## 5 Conclusions

Our findings supported a bidirectional relationship between sleep duration and cognitive function. We hypothesize short or long sleep duration was not only an unhealthy lifestyle, but also a result of the cognition-related neurodegenerative progress. For people with short or long sleep duration, their cognitive function might have already declined. A sufficient sleep duration is always recommended to achieve health aging. More attention should be paid on the cognition of those people. Cognitive training is recommended^52,53^, which might improve sleep duration due to our theory.

## Data Availability

The data used in this manuscript from the China Health and retirement Longitudinal Study (CHARLS). We applied the permission for the data access (https://charls.pku.edu.cn/zh-CN) and got the access to use it. Prof. Yaohui Zhao (National School of Development of Peking University), John Strauss (University of Southern California), and Gonghuan Yang (Chinese Center for Disease Control and Prevention) are the principle investigators.

## 6 Author contributions

Jianian Hua contributed conception and design of the study. Hongpeng Sun and Jianian Hua organized the database. Hongpeng Sun and Jianian Hua performed the statistical analysis. Hongpeng Sun and Jianian Hua wrote the first draft of the manuscript. Qi Fang reviewed the manuscript. All authors approved the final version of the paper.

## 7 Acknowledgments

We appreciated the China Center for Economic Research, the National School of Development of Peking University for providing the data.

## 8 Conflicts of interest

The authors declare that the research was conducted in the absence of any commercial or financial relationships that could be construed as a potential conflict of interest.

## 9 Funding

This study is supported by grants from the National Key Research & Development Program of China (No. 2017YEE0103700) and General Program of National Natural Science Foundation of China (No. 82071300).

## 10 Ethics statement

Ethical approval for all the CHARLS waves was granted from the Institutional Review Board at Peking University. The IRB approval number for the main household survey, including anthropometrics, is IRB00001052-11015; the IRB approval number for biomarker collection, was IRB00001052-11014. During the fieldwork, each respondent who agreed to participate in the survey was asked to sign two copies of the informed consent, and one copy was kept in the CHARLS office, which was also scanned and saved in PDF format. Four separate consents were obtained: one for the main fieldwork, one for the non-blood biomarkers and one for the taking of the blood samples, and another for storage of blood for future analyses.

## Notes

### Competing Interest Statement

The authors have declared no competing interest.

### Clinical Trial

This is a retrospective observational cohort study. The detail of this study was described in the article "Cohort profile: the China Health and Retirement Longitudinal Study (CHARLS)", doi:10.1093/ije/dys203

### Author Declarations

Ethical approval for all the China Health And Retirement Longitudinal Study (CHARLS) waves was granted from the Institutional Review Board at Peking University. The IRB approval number for the main household survey, including anthropometrics, is IRB00001052-11015; the IRB approval number for biomarker collection, was IRB00001052-11014. During the fieldwork, each respondent who agreed to participate in the survey was asked to sign two copies of the informed consent, and one copy was kept in the CHARLS office, which was also scanned and saved in PDF format. Four separate consents were obtained: one for the main fieldwork, one for the non-blood biomarkers and one for the taking of the blood samples, and another for storage of blood for future analyses.

