## Supplemental Material for "Bidirectional Associations between Short or Long Sleep Duration and Cognitive Function: the China Health and Retirement Longitudinal Study"

### 1 Tables

#### 1.1 Table S1. Associations between the trajectories of the combined latent growth models

|  | r | P value |
| --- | --- | --- |
| S intercept-S slope | 0.000 (0.000) | 0.090 |
| GC intercept-GC slope | 0.267 (0.028) | <0.001 |
| S intercept- GC intercept | -0.120 (0.009) | <0.001 |
| S slope-GC slope | -0.001 (0.000) | 0.059 |
| S intercept-GC slope | 0.001 (0.001) | 0.066 |
| GC intercept-S slope | -0.003 (0.002) | 0.079 |

Abbreviations: S, sleep duration; GC, global cognition.

#### 1.2 Table S2. Unstandardized coefficients of confounders on trajectories of sleep duration and cognitive function

|  | Sleep duration  baseline | Sleep duration  slope | Global cognition  baseline | Global cognition  slope |
| --- | --- | --- | --- | --- |
| Age | 0.003^***^ | <0.001^**^ | -0.102^***^ | -0.012^***^ |
| Sex | -0.04^***^ | -0.005^***^ | 1.839^***^ | 0.001 |
| Living area | -0.028^***^ | -0.003^*^ | 2.671^***^ | 0.039^*^ |

^***^P<0.001

#### 1.3 Table S3. Confounders in cross-lagged models which across all four waves

|  | Age | Sex | Living area |
| --- | --- | --- | --- |
| Standardized |  |  |  |
| Sleep duration Wave 2 | 0.025 | 0.002 | -0.055*** |
| Sleep duration Wave 3 | -0.004*** | -0.005*** | -0.003*** |
| Sleep duration Wave 4 | 0.050*** | 0.033* | -0.042** |
| Global cognition Wave 2 | -0.124*** | 0.090*** | 0.072*** |
| Global cognition Wave 3 | -0.103*** | 0.046*** | 0.084*** |
| Global cognition Wave 4 | -0.105*** | 0.023*** | 0.041*** |
| Unstandardized |  |  |  |
| Sleep duration Wave 2 | 0.003 | 0.004 | -0.160*** |
| Sleep duration Wave 3 | -0.001*** | -0.011*** | -0.010*** |
| Sleep duration Wave 4 | 0.007*** | 0.080* | -0.127** |
| Global cognition Wave 2 | -0.046*** | 0.564*** | 0.570*** |
| Global cognition Wave 3 | -0.048*** | 0.360*** | 0.825*** |
| Global cognition Wave 4 | -0.061*** | 0.221** | 0.502*** |

^*^P<0.05, ^**^P<0.01, ^***^P<0.001.

#### 1.4 Table S4. Cross-lagged models which across two waves^a^

|  | Unstandardized β (SE) | Standardized β (SE) |
| --- | --- | --- |
| Sleep duration to global cognition | -0.359 (0.059) | -0.086 (0.014) |
| Global cognition to sleep duration | -0.026 (0.005) | -0.103 (0.019) |

^a^This model included Wave 1 and Wave 3. 4 h and 10 h were the cut-off points. AFI=1.000, TLI=1.000, RMSEA=0.002.

#### 1.5 Table S5. Confounders in cross-lagged models which across two waves^a^

|  | Age | Sex | Living area |
| --- | --- | --- | --- |
| Standardized |  |  |  |
| Sleep duration Wave 1 | -0.050*** | 0.002 | -0.026* |
| Sleep duration Wave 3 | 0.037** | -0.005 | -0.035** |
| Global cognition Wave 1 | -0.253*** | 0.236*** | 0.269*** |
| Global cognition Wave 3 | -0.191*** | 0.090*** | 0.116*** |
| Unstandardized |  |  |  |
| Sleep duration Wave 1 | -0.006*** | 0.005 | -0.065* |
| Sleep duration Wave 3 | 0.004** | -0.010 | -0.089** |
| Global cognition Wave 1 | -03119*** | 1.936*** | 2.774*** |
| Global cognition Wave 3 | -0.094*** | 0.770*** | 1.265*** |

^*^P<0.05, ^**^P<0.01, ^***^P<0.001.

^a^This model included Wave 1 and Wave 3.

### 2 Figures

**Figure S1. Association between changes in sleep duration and cognitive function.** Figure Reply S1(b) shows the Table 3 of *Zhu 2020*^1^*.* Figure S1(c) shows our findings^2^. Together with the two findings, our previous theory hypothesized a new association between changes in sleep duration and cognitive function: changes from short or long sleep duration to moderate sleep duration benefits cognition; deviations from the moderate sleep duration or “excessive change” harms cognition.

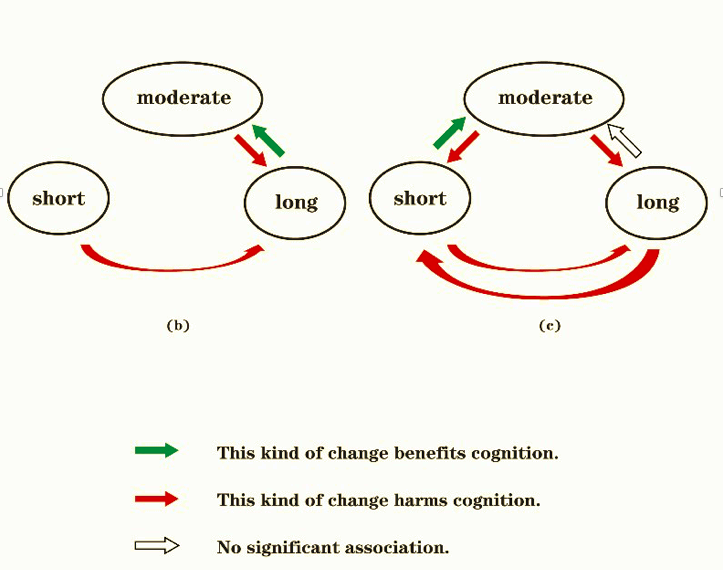

**Figure S2. Latent growth models of sleep duration and global cognition.**

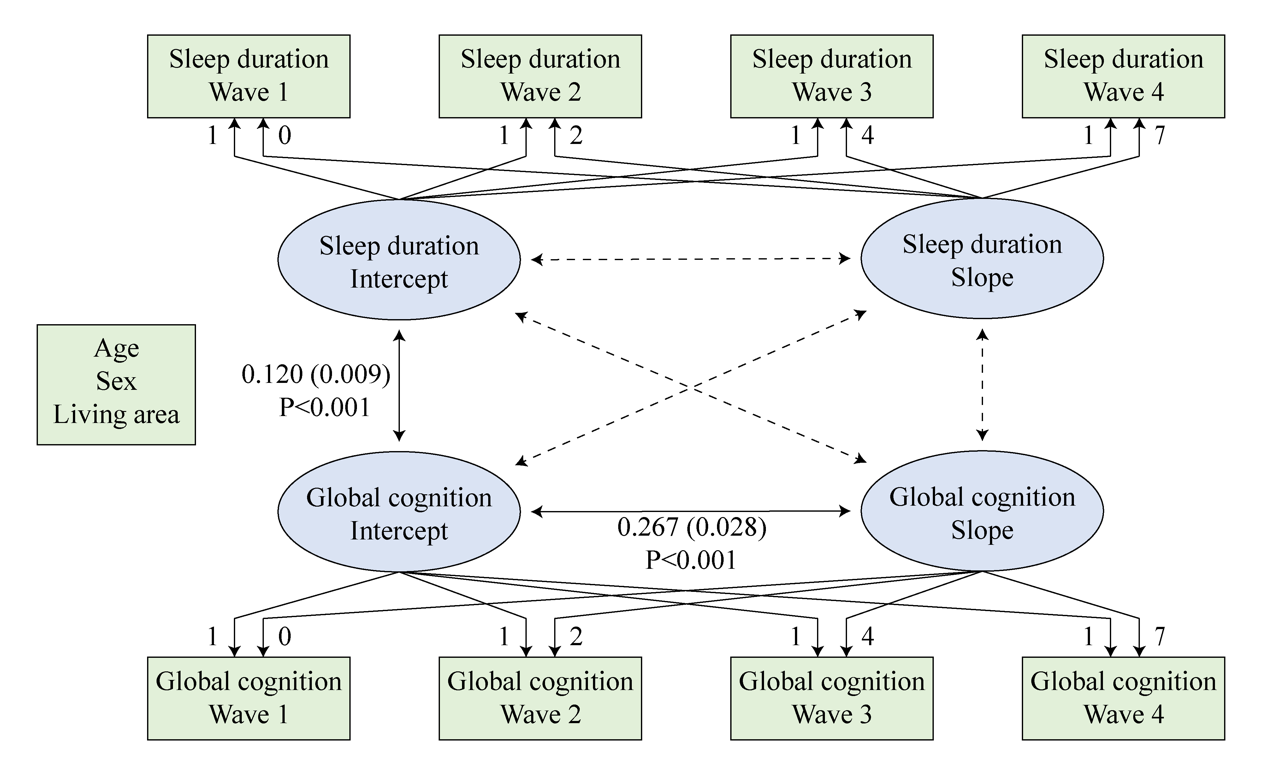
